# Image guidance improves freedom from recurrence in superficial radiation therapy for non-melanoma skin cancer

**DOI:** 10.1101/2022.09.29.22280478

**Authors:** Erin McClure, Geoffrey Sedor, Mairead Moloney, Yuxuan Jin, Michael W. Kattan, Lio Yu

## Abstract

**Importance:** This is the first study to quantify the 2-year recurrence rate of non-melanoma skin cancers (NMSCs) treated with image-guided superficial radiation therapy (IGSRT) and compare it to existing data on SRT, which is key to demonstrating the efficacy of IGSRT.

**Objective:** To quantify the 2-year recurrence rate for individuals with basal cell carcinoma (BCC), squamous cell carcinoma (SCC), and squamous carcinoma in-situ (SCCIS) treated with IGSRT versus SRT without image guidance.

**Design:** A retrospective cohort study that collected data from a standardized electronic medical record (Modernizing Medicine – EMA), written radiation treatment records, and record/verify system (Sensus Healthcare – Sentinel) to evaluate the 2-year recurrence rate of NMSCs treated by IGSRT (March 2016 to January 2022) and compare it to existing data on NMSCs treated by SRT via one sample proportion tests.

**Setting:** Multi-institution outpatient dermatology practices.

**Participants:** Individuals > 18 years old with biopsy-proven SCC, SCCIS, and/or BCC treated with IGSRT. 1602 patients with a total of 2,880 treated lesions were followed until 1/14/2022. As of that date, 22 lesions had recurred, and 2858 lesions were still at risk for recurrence. An 18-month cutoff for recency of assessment was utilized, resulting in 1204 lesions (41.8%) that were lost to follow-up.

**Exposure:** Treatment with IGSRT or SRT.

**Main Outcomes:** The overall 2-year recurrence probability of 2880 NMSCs treated by IGSRT was 0.7%, which is statistically significantly lower than two previously reported SRT studies (p *<* 0.001).

**Results:** All NMSCs (SCC, SCCIS, BCC) treated by IGSRT in this cohort had an aggregate 2-year freedom from recurrence of 99.23%. When stratified by NMSC histologic type, the recurrence rate for BCC was 1.1%(N=1382), for SCC 0.8% (N=904) and for SCC 0.0% (N=594). These rates of recurrence are significantly improved compared to a pooled study that evaluated NMSCs across histology (Cognetta) and BCCs alone (Silverman) treated without image guidance (standard SRT) (p < 0.001).

**Conclusions and relevance:** Image-guided superficial radiation (IGSRT) therapy offers a paradigm-shifting treatment option for patients with NMSCs – offering statistically significantly improved outcomes compared to standard SRT, and a more desirable toxicity profile to surgical options. This study demonstrates that this treatment modality is associated with remarkably low recurrence rates, which are statistically significantly improved from the previous generation of SRT, and in line with modern outcomes for Mohs micrographic surgery, though a head-to-head comparison has yet to be performed.

**Key Points:** *Question:* Does treatment by new generation image-guided superficial radiation therapy (IGSRT) improve the rates of recurrence of nonmelanoma skin cancers (NMSCs) compared to non-image-guided SRT?

*Findings:* This cohort study evaluated the recurrence rates of 2880 NMSCs treated with IGSRT. In this dataset, the 2-year rate of recurrence for IGSRT-treated NMSCs collectively was 0.7%. This is statistically significantly lower than the recurrence rates of NMSC treated by SRT without image guidance (1.9% Cognetta; 6.3% Silverman).

*Meaning:* Recurrence rates in NMSCs when treated with IGSRT are statistically significantly improved from SRT without image guidance, supporting the use of this new technology in clinical practice.

## Introduction

Non-melanoma skin cancers (NMSCs) are primarily comprised of basal cell carcinomas (BCC) and squamous cell carcinomas (SCC).^1^ While they are not nearly as fatal as melanoma, they can be a source of significant morbidity and they are the most common malignancy found in the United States.^2^ Unfortunately, their incidence is rising in many areas of the world, including the USA^2^–4, where incidence is increasing by approximately 2% annually.^5^ The incidence of NMSCs in the USA was estimated to be 5.4 million cases in 2012, which is a 35% increase since 2006.^6^

Currently, the primary treatment modality for low risk NMSCs is surgical resection.^1^ Patients with advanced disease are often treated with systemic therapies and in clinical trials or with comprehensive radiation therapy. For patients with localized disease, but that are poor surgical candidates (comorbidities/advanced age, inoperable location, morbid surgical outcomes, etc.) definitive radiation therapy is preferred. Definitive dosing tends to range from 30 – 60 Gy.^7^ Radiation therapy can also be useful as adjuvant treatment, especially when there are close or positive surgical margins. Additionally, it can act as a palliative option in the setting of advanced disease.^8^

Superficial radiation therapy (SRT) is a type of external radiation therapy that utilizes a lower level of energy which limits penetration to tissues beyond the skin. Dosing schedules for SRT typically vary from 5 Gy/fraction x 7 fractions (35 Gy total) to 2 Gy/fraction x 30 fractions (60 Gy total) depending on age of the patient, lesion location, and lesion size.^9^ This modality has been in use for over a century and was commonly practiced in the 1970s, with up to 55% of dermatology clinics utilized SRT in 1974.^7^ SRT was phased out as the preferred modality secondary to the advent of Mohs micrographic surgery (MMS), which has a superior recurrence-free survival to SRT (MMS has 99% 5-year recurrence-free survival for BCC and 97% for SCC versus SRT which has 96% 5-year recurrence-free survival for BCC and 94% for SCC).^10,11^

IGSRT employs the use of ultrasound technology to better visualize the cancerous lesion, which allows for more precise targeting of the radiation by accurately assessing the tumor margin and determining the necessary depth of treatment. Specifically, an ultrasound set to a frequency of 22 MHz (optimal for assessing the skin layer at a depth of 0-6mm) is used to determine the extent of the lesion beyond what is clinically visible.^12^ We hypothesize that the improved NMSC visualization due to the use of ultrasound to guide SRT will improve the 2-year freedom from recurrence rates of these lesions. Data supporting this hypothesis would shift the paradigm of radiation therapy for the use of NMSCs and help make it a more viable option for certain patients that are poor candidates for MMS, or for whom the toxicity profile is more favorable.

## Methods and Data

Treatments with IGSRT were initiated between 3/28/2016 and 1/6/2020. The initial dataset was obtained from a published article which had 2917 lesions, however 19 lesions had no follow-up after treatment completion, 14 lesions had mixed histologies, and 4 lesions had no histology recorded, so these lesions were excluded.^12^ Patients were followed until 1/14/2022. On that date, 22 lesions had recurred, and 2858 lesions were still at risk for recurrence. Using an 18-month cutoff for recency of assessment, 1204 lesions (41.8%) were lost to follow up (i.e., had not been assessed after 7/14/2020). The median follow-up among patients who remain alive and did not recur was 26.3 months (1st and 3rd quartiles, 10.0 and 38.4 months, respectively) with a maximum of 50.1 months.

Time to lesion recurrence was estimated using the cumulative incidence method with death of the host as a competing risk. Observed 2-year probabilities were compared with those of the literature with a test of proportions.

### Pooled Analysis Method

In addition to individual comparisons by histology of reported groups, we desired to compute an overall, pooled outcome comparison. In order to conduct this comparison of the IGSRT cohort to those reported in the literature, we pooled results from two reference groups which reported enough granularity of outcome to compute 2-year freedom from recurrence rates. To do so, we summed the total number of events and total number of patients for the combined cohort for comparison to ours. A similar approach was used to determine recurrence rate for all patients in the reference groups across with BCC (which was included in both published groups cohorts). The IGSRT recurrence probabilities and these pooled estimates were compared using a test of proportions.

## Results

### Patient characteristics

The median age of patients at their first treatment with IGSRT was 74 (1st and 3rd quartiles 67 and 80). Twenty two patients (0.76%) experienced a recurrence of their NMSC and 70 patients died of causes unrelated to their NMSC. Median follow up was 26.3 months (1st and 3rd quartiles 10 to 38 months). See **Table 1** for further details.

**Table 1.** Cohort characteristics.

| Characteristic |  |
| --- | --- |
| Lesions | 2,880 |
| Patients | 1,602 |
| Age* | 74 (67, 80) |
| Gender |  |
| Male | 913 (57.0%) |
| Female | 689 (43.0%) |
| Histology |  |
| BCC | 1,382 (48.0%) |
| SCC | 904 (31.4%) |
| SCCIS | 594 (20.6%) |
| Stage |  |
| 0 | 594 (21%) |
| 1 | 1,896 (66%) |
| 2 | 390 (14%) |
| Follow up (months)* | 26.3 (10.0, 38.4) |
| Event |  |
| Death (other cause) | 70 |
| Recurrence | 22 |
| Total Dose (Gy) † | 52.2 (37.2, 73.6) |
| Treatments† | 20.1 (20, 30) |

\*Median (IQR); n (%); †Mean (min, max)

### Tumor characteristics and staging

The majority of NMSCs were BCCs at 48.0% (N=1382), with SCCs comprising 31.4% (N=904), and the 20.6% (N=594) were SCCis. BCCs were also the most likely to recur with a recurrence rate of 1.1%, versus 0.8% for SCC and 0.0% for SCCis. Most NMSCs were found on the head/neck (66%), where cosmesis becomes ever more important. The vast majority of NMSCs were stage I (66%), and stage 0 for SCCis (14%). The full clinical details can be found in **Supplemental Table 1 and 2** describing stage, and event type for each histology in our study. Adverse events have been reported previously,^12^ but overall this treatment is exceptionally well tolerated: in brief, using the Radiation Therapy Oncology Group (RTOG) skin toxicity grading system, of the 2154 lesions with documented RTOG grading, 79% (n=1698) were grade 1, 20% (n=436) were grade 2, and 0.9% were grade 3+.

### Outcomes

In this study’s cohort of 2880 lesions of 1602 patients treated primarily with IGSRT the 2-year recurrence rate of overall NMSC lesion recurrence was 0.7% (N =2880), see **Figure 1, top**. When separated by histology, BCC had a 2-year recurrence rate of 1.1% (N=1382), SCC was 0.8% (N=904), and SCCIS was 0.0% (N=594), see **Figure 1, middle**. This contrasts with the Cognetta study which evaluated 1715 lesions treated by SRT and had an overall 2-year NMSC recurrence rate of 1.9%, more than double that of the cohort studied here.^11^ By histology, Cognetta reported 2.0% for BCCs, 1.8% for SCCs and 1.9% for SCCISs. Lastly, the Silverman study which also utilized SRT, had a 2-year recurrence rate of 6.3% for BCCs.^13^

**Figure 1.**
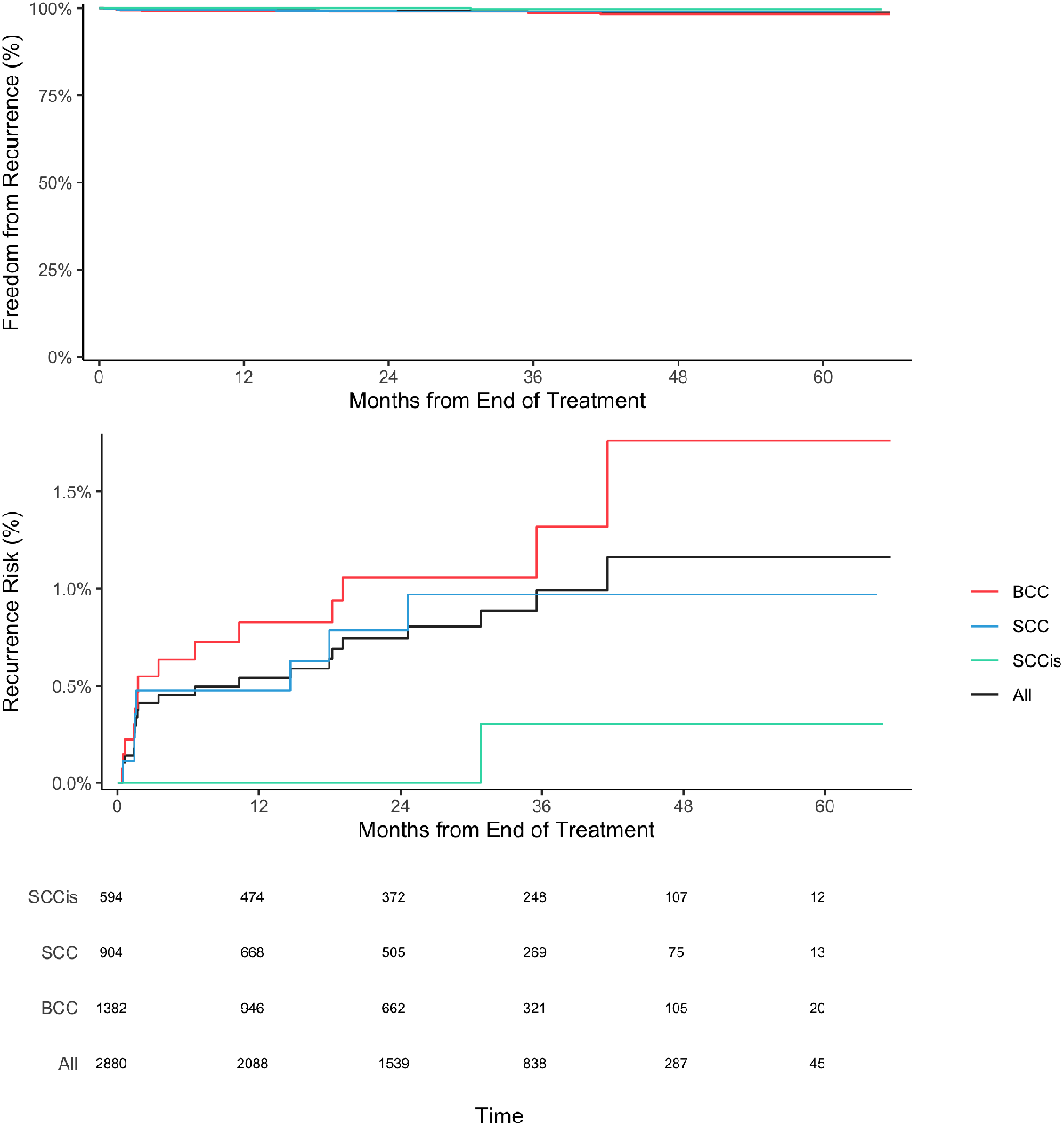
Freedom from recurrence over time. Top: Freedom from recurrence for all evaluated NMSCs (SCC, SCCIS, BCC). **Middle:** Cumulative incidence of recurrence for SCC (green), SCCIS (blue), BCC (red) and all histologies (black). **Bottom:** Number of evaluable patients over time.

### Comparison of recurrence rates

In the most general analysis, we compared recurrence data from all histologies in our IGSRT cohort directly to pooled data from the Cognetta and Silverman SRT studies together. Comparing these two pooled groups’ 2 year freedom from recurrence revealed a significantly improved outcome for the patients treated with IGSRT (p <0.001).

As a secondary analysis, we compared the two traditional SRT studies and our own data, subdivided by histology. All secondary histologic head to head comparisons show statistically significant differences favoring IGSRT: SCCis (p =0.0012), BCC (p =0.0163) and SCC (p =0.03) lesions as reported by Cognetta; and BCC as reported Silverman (p <0.001), see **Figure 2**.

**Figure 2.**
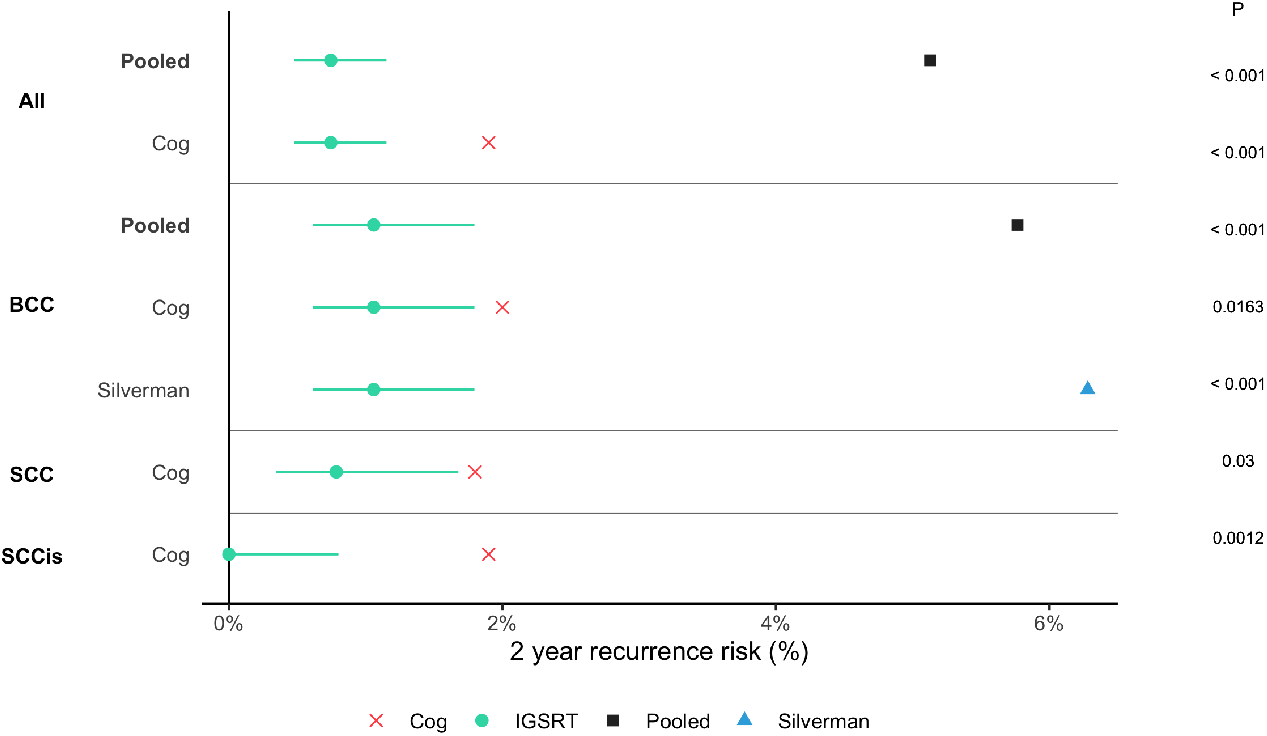
Forest plot of NMSC 2-year freedom from recurrence, IGSRT vs SRT. Circles represent median recurrence rate for our IGSRT cohort, and lines represent confidence intervals. Red X’s represent data from Cognetta study,^11^ and blue diamond from Silverman.^13^ Black squares represent pooled SRT cohorts for comparison.

## Discussion

### Major findings

The major findings of this study are the statistically significantly improved 2-year recurrence rates of NMSC when treated primarily with IGSRT compared to SRT. This held true for every comparison made between IGSRT and SRT (pooled and stratified by histology). This improved recurrence rate with IGSRT indicates that radiation may be a viable therapeutic option for patients with early-stage NMSCs like BCC, SCC, and SCCIS or those who cannot tolerate or are ill-suited for surgical resection.

### Risk of secondary cancer

The risk of developing a secondary cancer from the use of radiation therapy is low and the lag time from therapy to incidence is long, with ranges reported from 2-40 years, though the average is 15. People who receive low integral doses (*<* 5 Gy) of radiation, and/or radiation at an increased age experience further minimized risk.^14^ Therefore, it is reasonable to infer that individuals treated with IGSRT are likely at minimal risk of developing a secondary malignancy given the extremely low integral doses of radiation using this modality and the high average age at the time of treatment. To our knowledge, there are no reported case of secondary malignancy from SRT.

### Adverse Reactions

IGSRT is remarkably well tolerated by patients, greater than 99% of lesions developed only RTOG grade 1 or 2 cutaneous toxicity (erythema, epilation, patchy desquamation, moderate edema and/or decreased sweating). All toxicities recovered fully, most within two weeks. Details on the adverse reaction in this cohort have been reported previously.^12^

### Limitations

A limitation of this study includes its retrospective design, as randomized controlled trials are the gold standard for treatment comparisons. Therefore, correlations can be assessed, but causation cannot be. Additionally, not enough time has elapsed yet to determine 5-year recurrence rates, which are a common end point in evaluating recurrences of NM-SCs. Lastly, beyond histology, cohort matching was unable to be performed in this analysis, which increases the possibility that confounding factors exist.

## Summary

In summary, the 2-year freedom from recurrence for individuals with NMSC treated with IGSRT was statistically significantly improved from those without image guidance, and is on par with that of modern surgical technique. This implies that IGSRT may be an effective and valuable treatment option for BCCs, SCCs, and SCCIS when patients are not good candidates for surgical removal, or when the toxicity profile of IGSRT is more desirable. Evaluating the risks of recurrence based on specific patient and tumor characteristics such as age, patient ethnicity, tumor size, and tumor site, and eventually genomics^15^ will help better characterize the patient populations that would receive the most benefit from IGSRT.

Patients experience excellent outcomes when their early-stage NMSCs are treated by IGSRT. Specifically, the minimal side effects and high satisfaction scores^16^ associated with the standard dosing regimen of IGSRT and the very low 2-year freedom from recurrence rates of NMSCs treated by this modality. Taken together, these data indicate that IGSRT is a critical technilogical step forward providing modern, MMS-scale local control of NMSCs with a very tolerable toxicity profile.

## Conclusion

To our knowledge, this cohort study is the first to directly evaluate and compare 2-year freedom from recurrence rates in NMSC lesions primarily treated with IGSRT versus SRT considering competing risks. We reported 2-year recurrence data for BCCs, SCCs, SCCISs, both separately and overall, for lesions treated primarily by IGSRT, and then compared this to existing data on the recurrence rates for these cancers treated by SRT. We found that overall, NMSCs that received IGSRT had a 2-year recurrence rate of 0.7%, which is statistically significantly less than those treated with SRT (p *<* 0.001). While no head-to-head trial data exist, this recurrence rate is on par with MMS and signifies that IGSRT is an excellent option for patients who cannot undergo surgical removal of their NMSCs or who desire a different toxicity profile.

## Supporting information

STROBE checklist

## Data Availability

All data produced in the present study are available upon reasonable request to the authors

## Supplemental Information

## Code and data availability

The clinical data and code used for analysis are available upon request.

## Additional tumor information

Our cohort included NMSC in many anatomical locations, for details see **Supplemental Table 1**. A subset analysis including site is planned in future work.

Staging and events by histology.

**Supplementary Table 1.** Tumor sites included in our cohort of patients treated with IGSRT, stratified by histology.

| Site | BCC, N = 1,382 | SCC, N = 904 | SCCis, N = 594 | Total, N = 2880* |
| --- | --- | --- | --- | --- |
| Head and Neck (H&N) | 1003 | 537 | 371 | 1911 |
| *H&N sublocation* |  |  |  |  |
| Ear | 121 | 95 | 54 | 270 |
| Scalp | 52 | 83 | 58 | 193 |
| Forehead | 104 | 73 | 74 | 251 |
| Temple | 37 | 19 | 12 | 68 |
| Forehead/Temple | 0 | 0 | 1 | 1 |
| Eyebrow | 5 | 2 | 4 | 11 |
| Eyelid | 24 | 2 | 1 | 27 |
| Nose | 350 | 78 | 46 | 474 |
| Cheek | 191 | 120 | 92 | 403 |
| Cutaneous Lip | 48 | 13 | 7 | 68 |
| Mucosal Lip | 3 | 13 | 4 | 20 |
| Chin | 6 | 0 | 0 | 6 |
| Jawline | 2 | 3 | 0 | 5 |
| Chin/Jawline | 1 | 0 | 0 | 1 |
| Neck | 59 | 35 | 18 | 112 |
| Other | 0 | 1 | 0 | 1 |
| Extremities | 164 | 304 | 181 | 649 |
| *Extremities sublocation* |  |  |  |  |
| Hand | 11 | 78 | 53 | 142 |
| Other | 153 | 226 | 128 | 507 |
| Shoulder | 60 | 15 | 10 | 85 |
| Trunk | 152 | 46 | 31 | 229 |
| *Trunk sublocation* |  |  |  |  |
| Chest | 45 | 26 | 10 | 81 |
| Back | 96 | 20 | 21 | 137 |
| Other | 11 | 0 | 0 | 11 |
| Penis | 0 | 1 | 0 | 1 |

\*5 lesion locations not recorded

**Supplementary Table 2.**
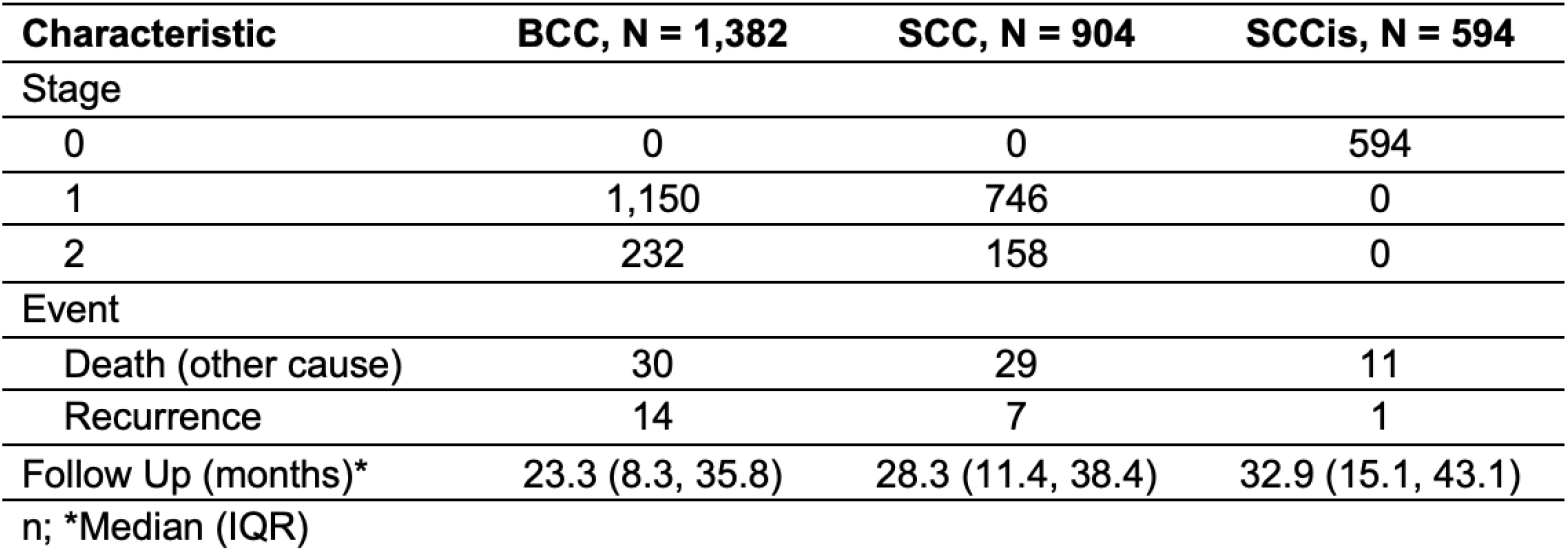
Clinical characteristics stratified by histology.

| Characteristic | BCC, N = 1,382 | SCC, N = 904 | SCCis, N = 594 |
| --- | --- | --- | --- |
| Stage |  |  |  |
| 0 | 0 | 0 | 594 |
| 1 | 1,150 | 746 | 0 |
| 2 | 232 | 158 | 0 |
| Event |  |  |  |
| Death (other cause) | 30 | 29 | 11 |
| Recurrence | 14 | 7 | 1 |
| Follow Up (months)* | 23.3 (8.3, 35.8) | 28.3 (11.4, 38.4) | 32.9 (15.1, 43.1) |

n; \*Median (IQR)

